# Mortality risk in survivors of acute COVID-19 and the urinary proteome: Follow-up results from a multinational study that prospectively evaluated a proteomic urine test for early and accurate prognosis of critical course complications in patients with SARS-CoV-2 infection (CRIT-COV-U)

**DOI:** 10.1101/2024.06.26.24309460

**Authors:** Justyna Siwy, Ralph Wendt, Felix Keller, Miroslaw Banasik, Björn Peters, Emmanuel Dudoignon, Alexandre Mebazaa, Dilara Gülmez, Goce Spasovski, Mercedes Salgueira Lazo, Harald Mischak, Manfred Hecking, Joachim Beige, UriCoV working group

## Abstract

Survival prospects following SARS-CoV-2 infection extend beyond the acute phase, influenced by various factors including age, health conditions, and infection severity. We investigated mortality risk among 651 post-acute COVID-19 patients, assessing the association between urinary peptides and future death. Data spanning until December 2023 were collected from six countries, comparing mortality trends with age- and sex-matched non-infected controls. A death prediction classifier was developed and validated using pre-existing urinary peptidomics datasets. Notably, 13.98% of post-COVID-19 patients succumbed during the follow-up, with mortality rates significantly higher than non-infected controls, particularly evident in younger individuals (<65 years). Urinary peptide analysis identified 201 peptides linked to mortality, integrated into a predictive classifier (DP201). Higher DP201 scores, alongside age and BMI, significantly predicted death. These findings underscore the utility of urinary peptides in prognosticating post-acute COVID-19 mortality, offering insights for targeted interventions.

## Main text

The life expectancy of SARS-CoV-2 survivors remains a subject of critical inquiry, affected by various factors such as age, pre-existing health conditions, and the severity of the initial infection^1^. While much attention has been directed towards understanding the acute phase of COVID-19, there is a growing recognition of the long-term health implications for those who have recovered. It has become increasingly evident that the impact of SARS-CoV-2 extends beyond the immediate symptoms experienced during the acute phase of the disease. The COVID-19 pandemic has markedly elevated global mortality^2^, with a significant proportion of COVID-19 survivors facing lingering health challenges and increased morbidity in the months and years following recovery. Among those at risk for severe outcomes, older individuals with pre-existing health conditions such as cardiovascular disease, diabetes, and respiratory disorders are particularly vulnerable^3,4^. Male sex, ethnicity (Black or South Asian), the severity of the initial infection have been found to correlate with Covid-19-related death^3^. However, long-term mortality among COVID-19 survivors is poorly understood, with specific underlying mechanisms remaining elusive. This study seeks to address this knowledge gap by examining mortality rates among patients who have survived the acute phase of the illness and by identifying specific urinary peptides that may be associated with future death.

By investigating mortality rates among patients who have survived the acute phase of COVID-19, we aim to identify potential predictors of long-term mortality, including demographic factors, clinical characteristics, and laboratory parameters. In a further approach, we focus on the role of urinary peptides as biomarkers of future death, leveraging recent advances in proteomic technology to explore their predictive value. Detection of urinary peptides associated with future mortality could have significant implications for risk stratification and personalized care among COVID-19 survivors. By incorporating these biomarkers into clinical practice, healthcare providers may be better equipped to identify individuals at high risk of adverse outcomes and to tailor interventions accordingly, to ultimately improve long-term outcomes and quality of life for survivors of COVID-19, contributing to more effective management of the ongoing pandemic.

Survival data of 651 unvaccinated patients were gathered from the “Prospective Validation of a Proteomic Urine Test for Early and Accurate Prognosis of Critical Course Complications in Patients with SARS-CoV-2 Infection” (CRIT-COV-U) study, spanning until December 2023 across sixcountries^5^. These patients were enrolled during the initial and subsequent waves of the pandemic in 2020-2021, predominantly infected with the wild-type virus, and had successfully passed the acute phase of COVID-19, defined as 21 days post-infection. The project complied with the Helsinki declaration. The Ethics Committee of the German-Saxonian Board of Physicians (Dresden, Germany; number EK-BR-70/23-1) and the Institutional Review Boards of the recruiting sites provided ethical approval. To assess the impact of age on mortality within this cohort, comparisons were made against age- and sex-matched data from non-SARS-CoV-2 infected individuals (n=5192), sourced from the human urinary database^6^.

Upon urine sampling, the mean age of the patients was 63 years (95%CI: 61-65), with a male predominance of 53.5%. The average body mass index (BMI) recorded was 27.0 (95%CI: 26.5-27.4) kg/m^2^, and the estimated glomerular filtration rate (eGFR) was at 90.0 (95%CI: 90.0-92.3) ml/min/1.73m^2^. The majority of patients, 56.4%, had no recorded comorbidities. A considerable portion of patients (47.8% of the cohort) experienced a relatively mild disease course (WHO-score 1-3). Throughout the follow-up period, spanning an average of 2.92 years (95%CI: 2.87-2.95), pertinent data was collected to assess mortality outcomes.

Among the 651 patients who survived the acute phase of COVID-19, 91 individuals (13.98%) succumbed during the follow-up duration, with 55 (8.45%) of these fatalities occurring within the initial year post-infection. In stark contrast, among the age and sex-matched controls totaling 5192 individuals, a markedly lower proportion of 92 (1.77%) deaths were recorded within the same timeframe. Notably, mortality displayed an age-dependent pattern across both cohorts, with significantly elevated rates observed among those who had survived COVID-19 compared to their non-infected counterparts (**Figure 1 A-D**). Specifically, within the first year post-infection, mortality rates surged up to 4.7 times higher in patients younger than 65 years compared to the non-infected controls.

**Figure 1:**
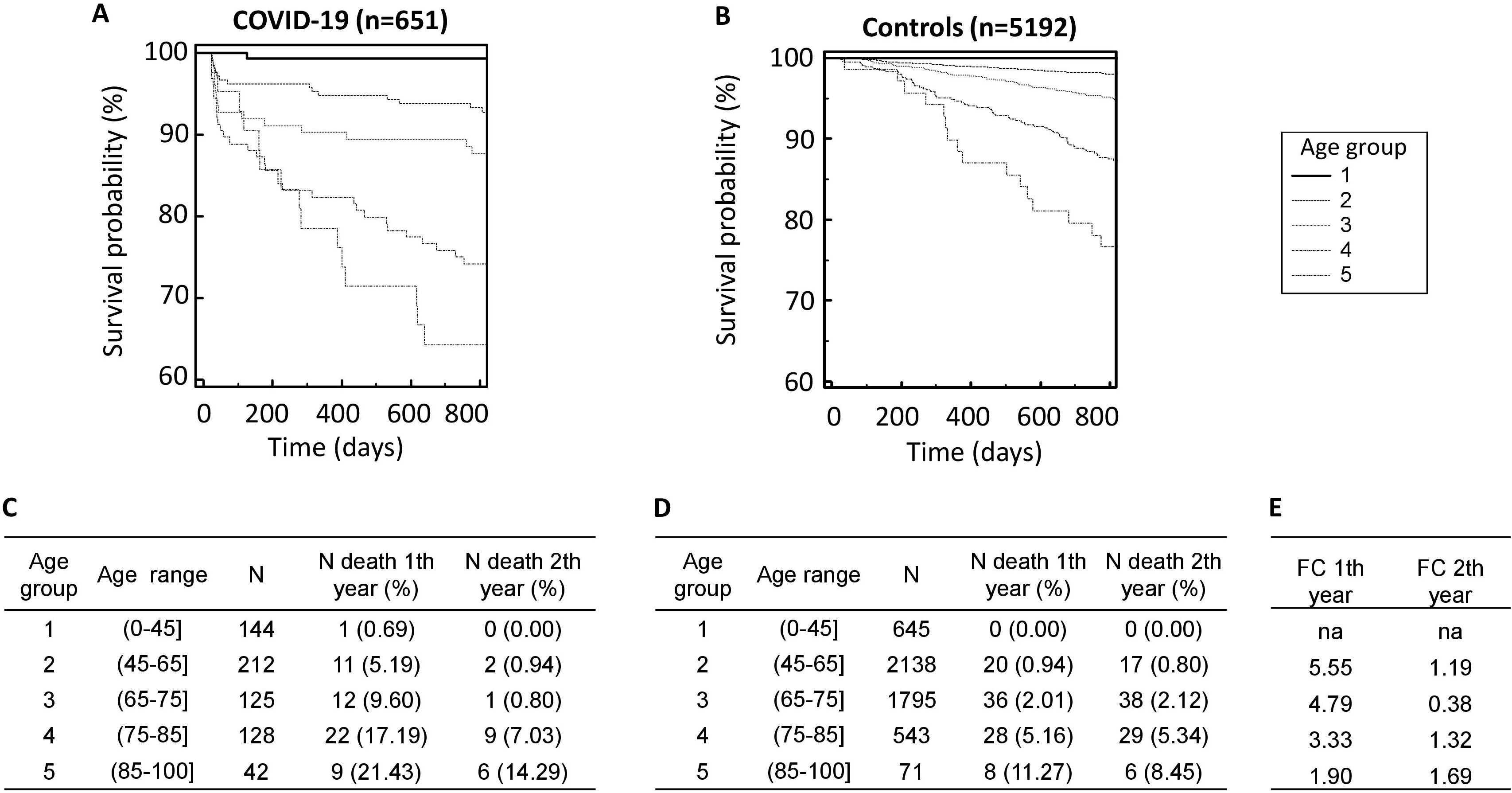
Age-dependent mortality in the COVID-19 cohort and healthy controls. Kaplan Meier survival curves for the cohort of survived acute phase COVID-19 cohort (A) and age and sex matched not infected healthy controls (B) are shown. The numbers of deaths per age group in the COVD-19 cohort (C) and controls (D) as well as the calculated fold change (FC) between the COVID-19 and controls (E) is given.

For the development and validation of a classifier aimed at predicting death after surviving the initial acute phase of COVID-19, pre-existing urinary peptidomics datasets from baseline samples of COVID-19-diagnosed patients within the CRIT-COV-U study were utilized. These datasets were stratified into training (n=324) and test (n=327) sets through random partitioning. The identification of peptides associated with mortality commenced by applying the Mann-Whitney test to compare 44 deceased and 280 surviving patients within the training set. Subsequently, adjustments for multiple testing were implemented to ensure statistical robustness. Independent validation of the identified peptides was then carried out using the test set, consisting of 47 deceased and 280 surviving patients.

The analysis of urinary peptidome datasets within the training set enabled the identification of 201 peptides (listed in supplementary **Table 1**) as significantly associated with mortality when comparing deceased and surviving patients. Among these peptides, 14 overlapped with the previously established Cov50 classifier designed for prognosticating unfavorable COVID-19 outcomes during the acute phase^7^. These 201 peptides were combined to form a support-vector-machine-based classifier (DP201) which was subsequently applied to the test cohort. The resultant outcomes are depicted in **Figure 2 (A-B)**, illustrating a clear correlation between higher classification scores and heightened mortality risk.

**Figure 2:**
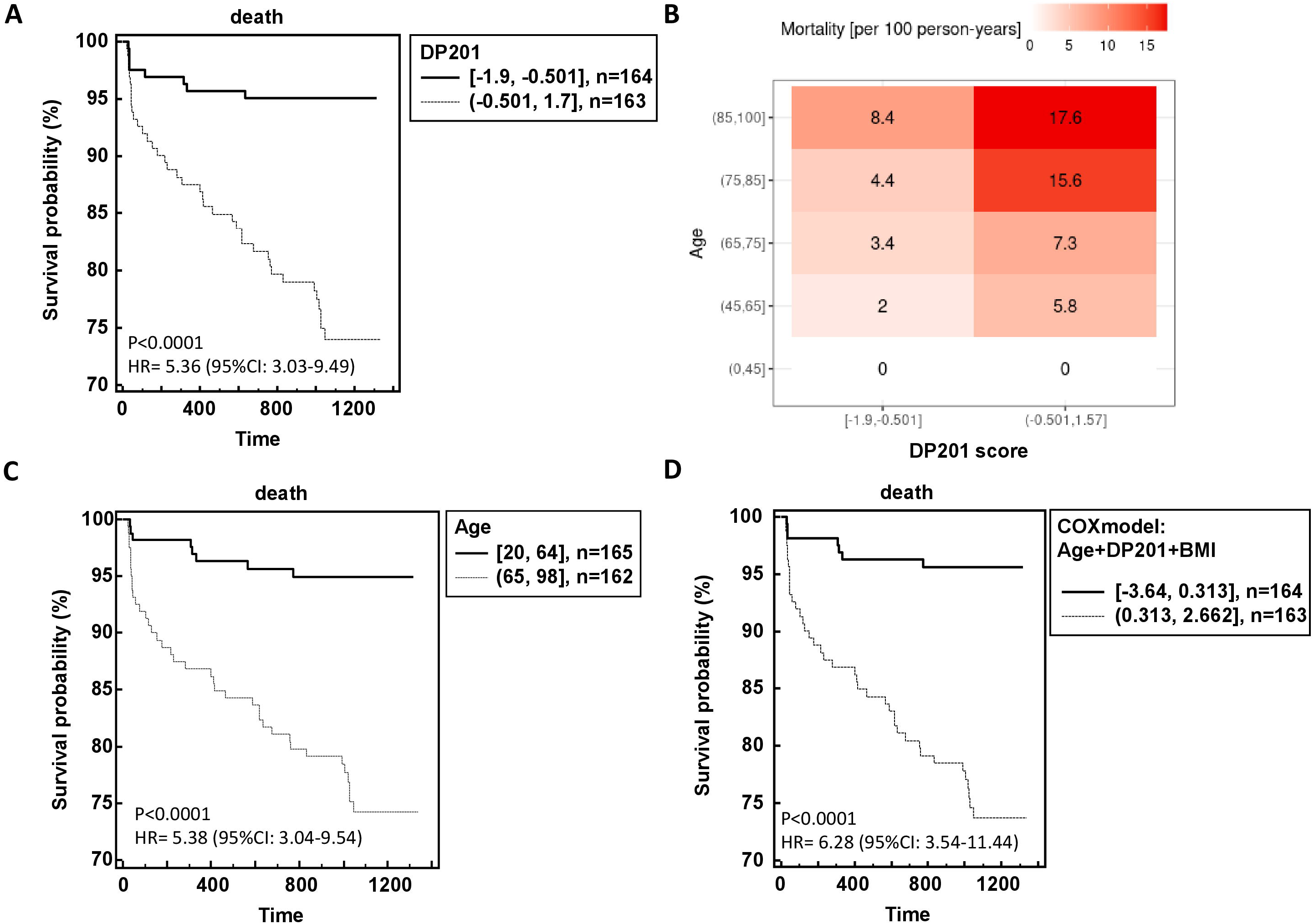
Performance of the urinary peptide-based death prediction classifier in the independent test data. The risk of death is significant dependent of the DP201 score (A) although the age dependency can still be observed (B). The hazard ration for survival probability DP201 classifier (A), and age (C) could be increased using a model including DP201, age and BMI (D).

Further Cox regression analysis revealed that age, BMI, and DP201 significantly influenced death prediction, while sex, number of comorbidities, eGFR, and WHO score did not exhibit statistical significance. Integration of these three parameters into a Cox model yielded a hazard ratio of 6.28 (95%CI: 3.54-11.44) compared to age and DP201 alone (**Figure 2 C-D**).

The peptides encompassed within the DP201 classifier comprised upregulated fragments of albumin, alpha-2-HS-glycoprotein, apolipoprotein A-I, and beta-2-microglobulin, alongside downregulated fragments of CD99 antigen, various collagens, fibrinogen alpha, polymeric immunoglobulin receptor, sodium/potassium-transporting ATPase, and uromodulin. These findings shed light on the molecular signatures associated with mortality risk among COVID-19 survivors, potentially paving the way for refined prognostic assessments and personalized interventions in clinical practice.

SARS-CoV-2 infection poses a persistent threat to individuals even beyond the acute phase, extending well beyond the initial 21-day period post-infection. Importantly, among those who successfully navigate the acute phase of COVID-19, the risk of mortality escalates significantly during the subsequent follow-up period. Particularly noteworthy is the observation that within the first year following infection, mortality rates surge dramatically among individuals who have survived the acute phase of the illness when compared to a non-infected control cohort. What is striking is that this increase in mortality risk is most pronounced among younger individuals, highlighting a concerning trend that defies conventional assumptions regarding age-related vulnerability to severe outcomes. Furthermore, the identification of specific urinary peptides capable of predicting heightened mortality risk at the outset of SARS-CoV-2 infection underscores the intricate interplay between molecular biomarkers and clinical outcomes. These peptides serve as early indicators of the likelihood of mortality, providing valuable insights into the underlying pathophysiological mechanisms driving adverse outcomes in COVID-19 patients. By leveraging these predictive biomarkers, healthcare professionals can proactively identify individuals at elevated risk of mortality and implement targeted interventions aimed at mitigating this risk, thereby potentially altering the trajectory of the disease course, like already showed for chronic kidney and heart diseases^8^.

In essence, the findings underscore the multifaceted nature of SARS-CoV-2 infection, extending far beyond the acute phase and exerting a lasting impact on mortality outcomes. By elucidating the dynamics of mortality risk in COVID-19 survivors and identifying predictive biomarkers, this research not only enhances our understanding of the long-term consequences of the disease but also lays the groundwork for more effective risk stratification and personalized interventions, ultimately contributing to improved clinical management and patient outcomes.

## Data Availability

The data that support the findings of this study are available upon request from the corresponding author

## Acknowledgment

**UriCoV Working Group:** Justyna Siwy, Mosaiques Diagnostics GmbH, Hannover, Germany; Ralph Wendt, Division of Nephrology, St. Georg Hospital, Leipzig, Germany; Joachim Beige, Division of Nephrology, St. Georg Hospital, Leipzig, Department of Internal Medicine II, Martin-Luther-University Halle-Wittenberg, Halle, Germany and Kuratorium for Dialysis and Transplantation (KfH) Leipzig, Leipzig, Germany, Miroslaw Banasik, Department of Nephrology and Transplantation Medicine, Wrocław Medical University, Wroclaw, Poland; Björn Peters, Department of Molecular and Clinical Medicine, Institute of Medicine, the Sahlgrenska Academy at University of Gothenburg, Gothenburg, Sweden and Department of Nephrology, Skaraborg Hospital, Skövde, Sweden; Emmanuel Dudoignon, Hospital Saint Louis-Lariboisière, Paris, France; Dilara Gülmez, Lenka Grula, Amelie Kurnikowski, Manfred Hecking, all from Department of Epidemiology, Medical University of Vienna, Vienna, Austria; Magdalena Krajewska, Justyna Zachciał, Dorota Bartoszek, Patryk Wawrzonkowski, Krzysztof Wiśnicki, from Department of Nephrology and Transplantation Medicine, Wrocław Medical University, Wroclaw, Poland; Emelie Sarenmalm, Region Västra Götaland, Skaraborg Hospital, Department of Infectious Diseases, Skövde, Sweden; Åsa Nilsson, Region Västra Götaland, Skaraborg Hospital, Research, Education, Development and Innovation Department, Skövde, Sweden; Goce Spasovski, University Sts. Cyril and Methodius, Skopje, Republic of North Macedonia; Mercedes Salgueira Lazo, Virgen Macarena Hospital and University of Seville, Sevill; Maria Isabel García Sánchez, Biobank Node at Virgen Macarena Hospital, Seville, integrated in the Spanish National Biobanks Network (PT23/00134); Marek W Rajzer from First Department of Cardiology, Interventional Electrocardiology and Arterial Hypertension, Jagiellonian University Medical College, Kraków, Poland; Beata Czerwieńska, from Department of Nephrology, Endocrinology and Metabolic Diseases, Medical University of Silesia, Katowice, Poland; Magdalena Dzitkowska – Zabielska from Faculty of Physical Education, Gdańsk University of Physical Education and Sport and Centre of Translational Medicine, Medical University of Gdansk, Gdansk, Poland; Lukasz Fulawka from Molecular Pathology Centre Cellgen, Wrocław, Poland; Elena Nowacki, University of Patients-Sorbonne University, Paris, France; Catherine Tourette-Turgis, University of Patients, research chair “Compétences & vulnérabilités” Sorbonne University, France; Morgane Michel, Université Paris Cité, ECEVE, UMR 1123, Inserm, Paris, France; Assistance Publique-Hôpitaux de Paris, Hôpital Robert Debré, Unité d’épidémiologie clinique, Paris, France.

